# Aldosterone after menopause: the Study of Women’s Health Across the Nation

**DOI:** 10.64898/2026.03.10.26348095

**Authors:** James Brian Byrd, Satish P. RamachandraRao, Shichi Dhar, Michelle M. Hood, Aleda M. Leis, Richard J. Auchus, Daniel McConnell, Samar R. El Khoudary, Rebecca C. Thurston, Carrie Karvonen-Gutierrez

## Abstract

**Background:** Hypertension (HTN) is a leading risk factor for mortality and disability, often as a result of heart disease or stroke, two leading causes of death. A blood pressure rise in midlife women in industrialized societies remains poorly understood. Progressively higher aldosterone levels have been associated with proportionately higher blood pressure in some populations. We examined whether aldosterone is associated with HTN in Study of Women’s Health Across the Nation (SWAN).

**Methods:** SWAN is a longitudinal cohort of women followed from midlife into late adulthood. Analyses include 999 women free of heart failure and with serum aldosterone measured during 2015-2016 (15th follow-up) study visit (V15). HTN was defined at V15 as systolic blood pressure (SBP) or diastolic blood pressure (DBP) ≥140 or ≥90mmHg, respectively, or use of antihypertensive medications. Utilizing the longitudinal data available in SWAN, treatment resistant hypertension (TRH) was defined as reported use of ≥4 concurrent antihypertensive medications at any visit, or ≥3 concurrent antihypertensive medications and SBP≥140 or DBP≥90 at two consecutive visits. Multivariable regression related aldosterone to HTN or TRH, adjusting for age, race/ethnicity, body mass index, physical activity, smoking, and low-density lipoprotein cholesterol.

**Results:** At V15, women were 66±2.7 years of age, and the prevalence of HTN and TRH was 52% and 4%, respectively. Each 1 ng/dL higher aldosterone was associated with a 4% increased odds of HTN (95%CI 1.02,1.06;p<0.001); association was not significant for TRH.

**Conclusions:** Our findings extend growing evidence that subclinical aldosterone excess is associated with greater HTN risk in postmenopausal women.

## INTRODUCTION

Hypertension (HTN) is a leading risk factor for heart disease or stroke, two leading causes of death.^1^ From 2017 to 2020, using the latest definition of hypertension (≥130/80 mm Hg), about 46.7% of adults aged ≥20 years in the United States (U.S.) had hypertension: 50.4% of males and 43.0% of females.^2^ Furthermore, serial cross-sectional analyses of participants in the National Health and Nutrition Examination Survey (NHANES) showed worsened control rates of high blood pressure, with the age-adjusted estimated proportion with controlled blood pressure (systolic BP <140 and diastolic BP < 90 mm Hg) decreasing from 53.8% in 2013-2014 to 43.7% in 2017-2018.^2^ Globally, the prevalences of hypertension and uncontrolled hypertension are high.^3^

The menopausal transition (MT) is a period of rapid endocrinologic and physiologic change. In that setting, some women experience a menopause-induced increase in blood pressure, distinct from an age-induced increase in blood pressure observed in other women.^4^ The physiological basis of these age-related and menopause-related changes remains poorly understood.

Dysregulation of the renin-angiotensin-aldosterone system (RAAS) in the presence of estrogen deficiency is a hypothesized contributor to blood pressure related changes in women during mid-life. 17β-estradiol has been found in animal model to suppress aldosterone production via actions on estrogen receptor β, but also stimulate aldosterone production via acitons on the G protein-coupled estrogen receptor.^5^ Clinical studies have documented that progressively higher aldosterone levels are associated with higher blood pressure as well as greater left ventricular remodeling and greater coronary artery calcification.^6^

Given the overall lack of information about the relationship between aldosterone and hypertension in aging women, our study tested the hypothesis that higher aldosterone is associated with higher blood pressure in the Study of Women’s Health Across the Nation (SWAN).^7^

## METHODS

### Study Population

The Study of Women’s Health Across the Nation (SWAN) is a longitudinal, multicenter study of the menopause transition and aging. The full sample consists of 3,302 women, aged 42-52 at the initial baseline visit completed in 1996. At baseline, these women were pre- or early perimenopausal, and self-identified with site-specific designated race and ethnic groups across seven locations: including White women (from all seven sites), Black women (from Boston, Massachusetts; Pittsburgh, Pennsylvania; Southeast Michigan; and Chicago, Illinois), Hispanic women (from Newark, New Jersey), Chinese women (from Oakland, California), and Japanese women (from Los Angeles, California). Of the 2,091 women (63.3% of the original cohort, 66.3% of the surviving cohort) who participated in the 2015-2016 follow-up visit [15th follow-up visit (V15)], the analysis utilized data from 1,014 women with serum aldosterone measurements available at V15. After excluding 15 women with a history of congestive heart failure by V15, the final analytic sample included 999 women (30.3% of the original cohort). Study protocols were approved by the Institutional Review Boards at each study site, and participants provided written, informed consent.

### Serum Aldosterone

The primary exposure measured was the level of circulating aldosterone (ALDO). Aldosterone levels were measured in sera samples using LC-MS/MS. The LC-MS/MS methods are published^8,9^ and described in further detail in the **Supplementary Methods**. The lower limit of quantitation (LLOQ) for the aldosterone assay was 2 ng/dL. Nearly one-quarter (n=238, 23.8%) of values were below the LLOQ and were imputed to be a random number between 0 and 2 ng/dL, using a random number generated from the uniform distribution in SAS v.9.4 (SAS Institute, Cary, NC). This allowed for the inclusion of participants whose aldosterone levels were below the LLOQ.

### Hypertension and Treatment-Resistant Hypertension

The two primary dependent variables were HTN and treatment-resistant hypertension (TRH). At each study visit from baseline through V15, blood pressure was measured by trained staff members using a standardized protocol and with a standard mercury sphygmomanometer or an automated sphygmomanometer and cuff placed on the participant’s right arm. Appropriate cuff size was determined based on arm circumference. Participants were seated and feet flat on the floor for at least five minutes prior to measurement. The average of two sequential measurements was used in analysis.

Pulse pressure was calculated as V15 systolic blood pressure (SBP) minus diastolic blood pressure (DBP). We defined hypertension at V15 as SBP or DBP ≥140 or ≥90 mmHg, respectively, or use of antihypertensive medications. This approach follows recent major epidemiologic studies and allows comparison with the extensive literature using this definition.^10^ In accordance with the American Heart Association’s Scientific Statement on resistant hypertension,^11^ treatment resistant hypertension (TRH) was defined utilizing the longitudinal data available in SWAN, as reported use of ≥4 concurrent antihypertensive medications at any visit, or ≥3 concurrent antihypertensive medications and SBP≥140 or DBP≥90 at two consecutive visits.

### Covariates

Covariates were collected through a combination of interviews and self-administered questionnaires. Participants provided self-reported information on their race and ethnicity, categorized as non-Hispanic White, Black, Chinese, Japanese, or Hispanic. With the exception of education, all other covariates were assessed at V15. Educational attainment was self-reported at study baseline and reported as less than a high school degree, high school degree, some college, college degree, or post-college. Level of economic strain reflected difficulties (very hard, somewhat hard, or not very hard) in paying for necessities including food, medical care, and housing. Smoking status was self-reported and categorized as current smoker, former smoker, or never smoker. Physical activity was assessed via a modified Baecke questionnaire,^12^ consisting of subscales for sports and exercise activity, nonsports leisure activity, and household and childcare activity, summed for a total score ranging from 3 to 15. Height (cm) and body weight (kg) were measured using a stadiometer and balance-beam scale, respectively, and used to compute body mass index (BMI, kg/m²). Triglycerides, total cholesterol, and high-density lipoprotein (HDL) were assayed at V15, and low-density lipoprotein (LDL) was calculated using the Friedewald formula (LDL = total cholesterol - HDL - triglycerides /five) when triglycerides were < 400 mg/dL.

### Statistical Analysis

Means and standard deviations (SD) were calculated for continuous variables and frequencies and percentages were calculated for categorical variables, overall and by V15 HTN status. Due to aldosterone being right-skewed, we calculated medians and interquartile ranges (IQRs) for aldosterone. Covariate data was compared across groups based on HTN status; differences were examined using t-tests for means, Kruskal-Wallis tests for medians, and Chi-square tests for categorical variables. Significant differences in medians of aldosterone by categorical covariates, including HTN and TRH were assessed using Kruskal-Wallis tests; significant pairwise associations were determined using Dunn’s test.

Then, logistic regression models examined relationships of each of HTN and TRH with aldosterone, adjusted for covariates in the following stages: 1) age, race and ethnicity, and BMI; 2) age, site, and BMI; 3) variables in the first stage plus smoking status and physical activity; and 4) variables in the third stage plus LDL. Logistic regression models of TRH utilized a Firth correction due to the small number of women with TRH. Finally, linear regression models of the relationship between SBP, DBP, and pulse pressure and aldosterone were examined, adjusting for all covariates except site, and including an interaction between aldosterone and antihypertensive medication use. Aldosterone was modeled as raw aldosterone value and log base 2-transformed aldosterone. In models including an interaction between antihypertensive medication use and continuous aldosterone, aldosterone was centered around its mean. Statistical significance was defined at α <0.05. All analyses were completed using RStudio version 2023.12.0.369.

## RESULTS

**Table 1** presents characteristics of the 999 women comprising the analytical sample. The mean age of the sample was 65.5 (±2.7) years (range: 60.4-72.2) at V15. Approximately half of the sample (51.4%) was non-Hispanic White, one quarter non-Hispanic Black (26.7%), 10.4% Chinese, 6.7% Japanese, and 4.8% Hispanic. The average BMI was 29.2 (±7.0) kg/m^2^. One-fifth reported economic strain at V15, and 50% did not have a college degree at baseline.

**Table 1.**
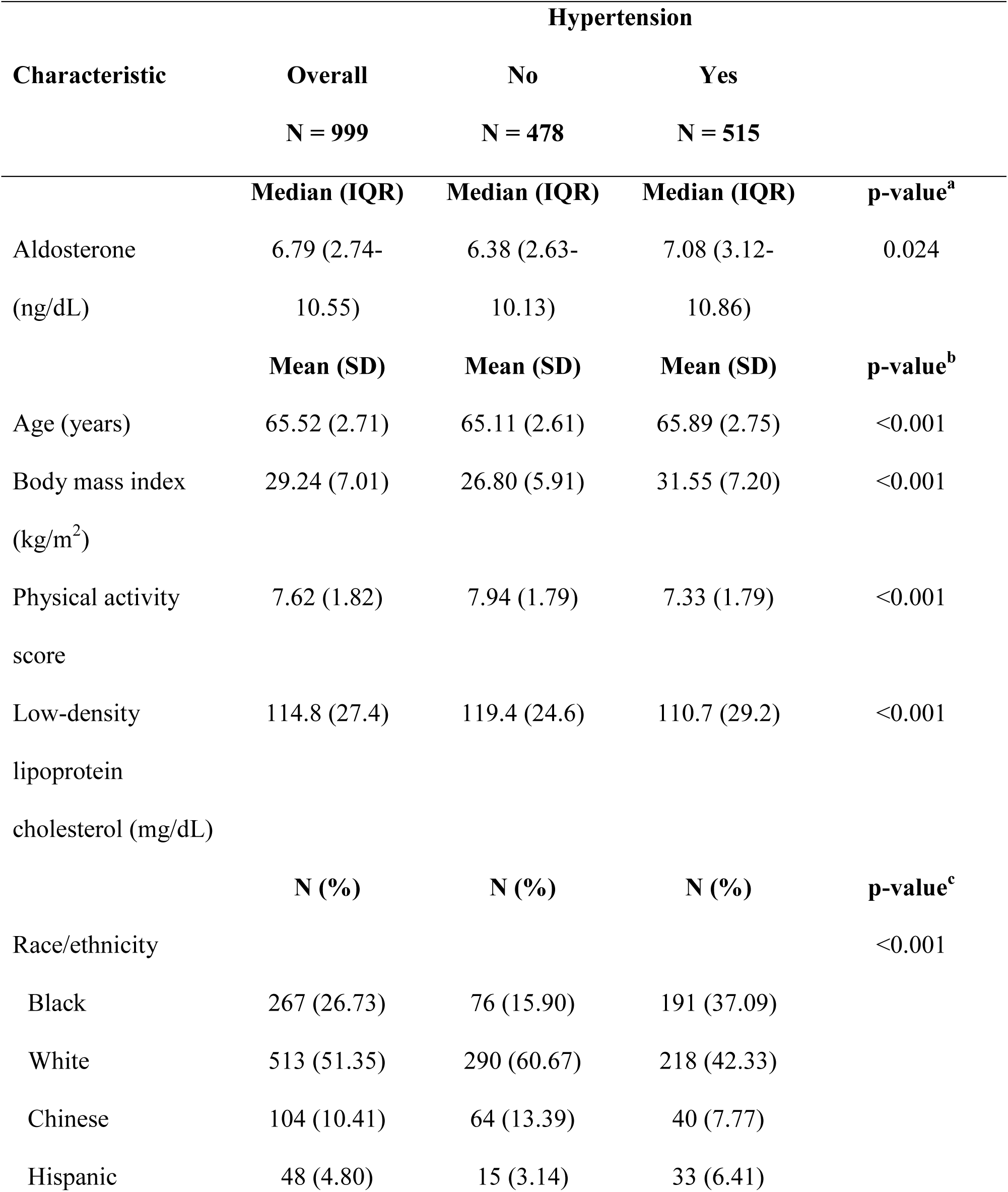
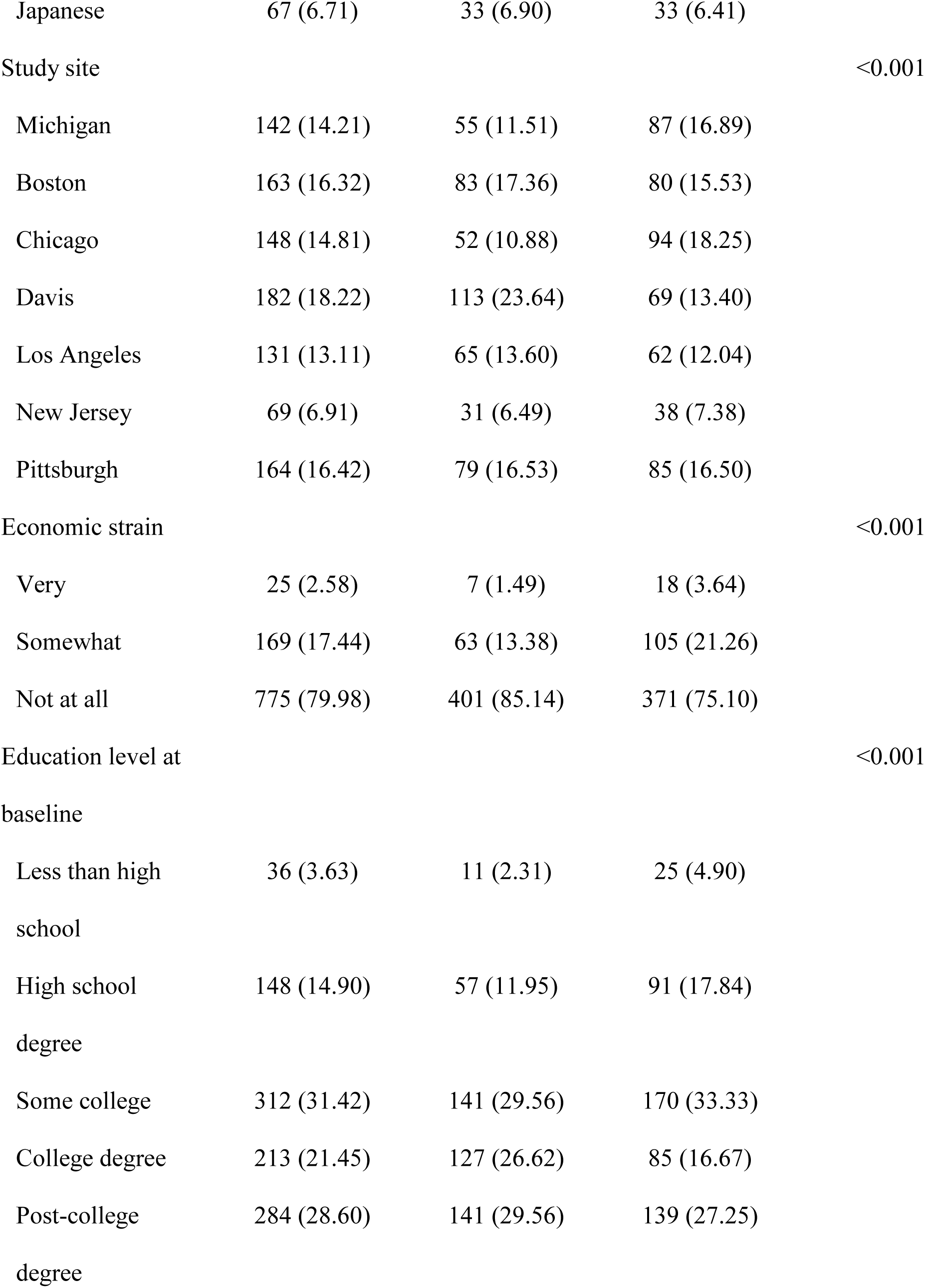
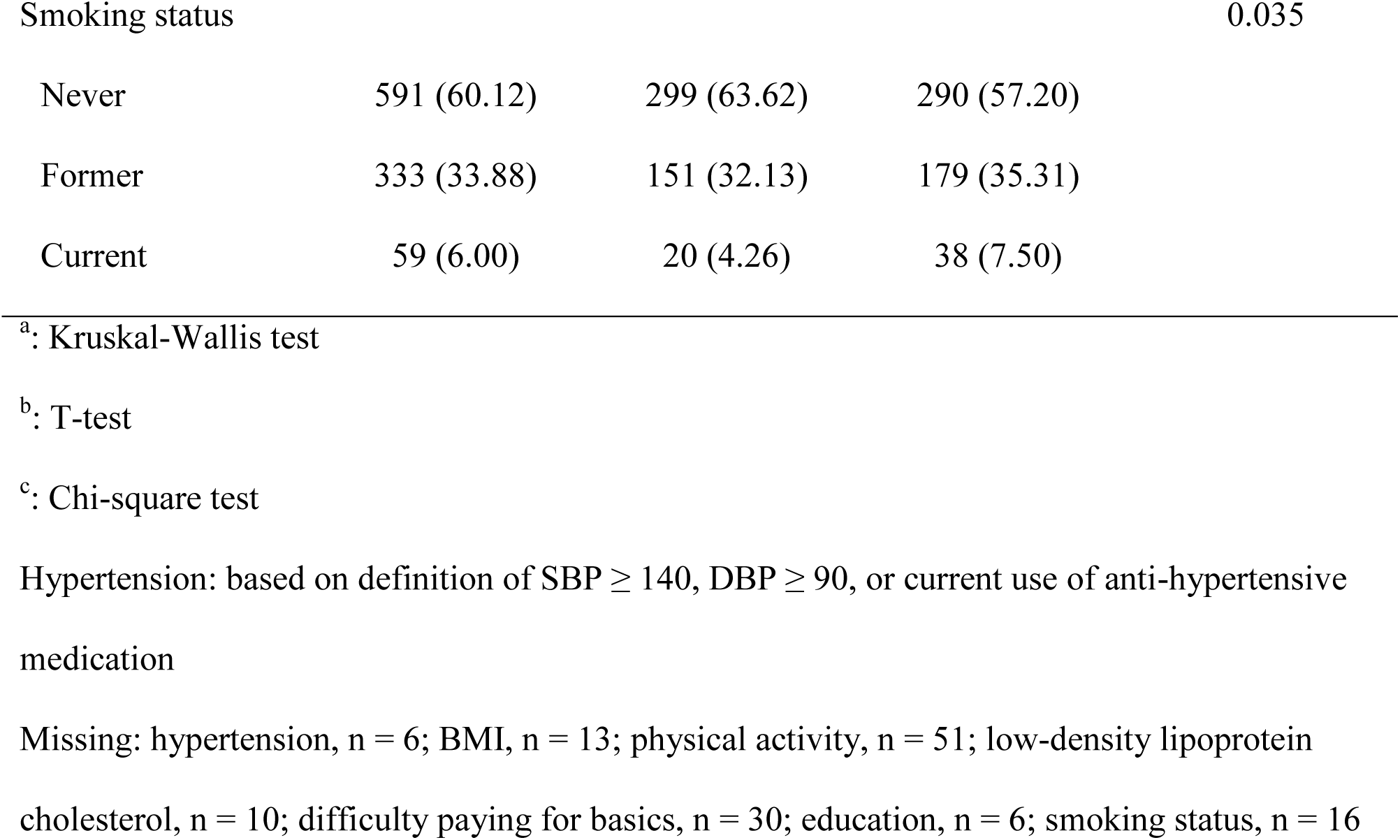
Population characteristics overall and by hypertension status at follow-up visit 15 (2015-16) in the Study of Women’s Health Across the Nation.

| Characteristic | Hypertension |  |  | p-value<sup>a</sup> |
| --- | --- | --- | --- | --- |
|  | Overall | No | Yes |  |
|  | N = 999 | N = 478 | N = 515 |  |
|  | Median (IQR) | Median (IQR) | Median (IQR) |  |
| Aldosterone<br>(ng/dL) | 6.79 (2.74-10.55) | 6.38 (2.63-10.13) | 7.08 (3.12-10.86) | 0.024 |
|  | Mean (SD) | Mean (SD) | Mean (SD) | p-value<sup>b</sup> |
| Age (years) | 65.52 (2.71) | 65.11 (2.61) | 65.89 (2.75) | <0.001 |
| Body mass index<br>(kg/m<sup>2</sup>) | 29.24 (7.01) | 26.80 (5.91) | 31.55 (7.20) | <0.001 |
| Physical activity<br>score | 7.62 (1.82) | 7.94 (1.79) | 7.33 (1.79) | <0.001 |
| Low-density<br>lipoprotein<br>cholesterol (mg/dL) | 114.8 (27.4) | 119.4 (24.6) | 110.7 (29.2) | <0.001 |
|  | N (%) | N (%) | N (%) | p-value<sup>c</sup> |
| Race/ethnicity |  |  |  | <0.001 |
| Black | 267 (26.73) | 76 (15.90) | 191 (37.09) |  |
| White | 513 (51.35) | 290 (60.67) | 218 (42.33) |  |
| Chinese | 104 (10.41) | 64 (13.39) | 40 (7.77) |  |
| Hispanic | 48 (4.80) | 15 (3.14) | 33 (6.41) |  |
| Japanese | 67 (6.71) | 33 (6.90) | 33 (6.41) |  |
| Study site |  |  |  | <0.001 |
| Michigan | 142 (14.21) | 55 (11.51) | 87 (16.89) |  |
| Boston | 163 (16.32) | 83 (17.36) | 80 (15.53) |  |
| Chicago | 148 (14.81) | 52 (10.88) | 94 (18.25) |  |
| Davis | 182 (18.22) | 113 (23.64) | 69 (13.40) |  |
| Los Angeles | 131 (13.11) | 65 (13.60) | 62 (12.04) |  |
| New Jersey | 69 (6.91) | 31 (6.49) | 38 (7.38) |  |
| Pittsburgh | 164 (16.42) | 79 (16.53) | 85 (16.50) |  |
| Economic strain |  |  |  | <0.001 |
| Very | 25 (2.58) | 7 (1.49) | 18 (3.64) |  |
| Somewhat | 169 (17.44) | 63 (13.38) | 105 (21.26) |  |
| Not at all | 775 (79.98) | 401 (85.14) | 371 (75.10) |  |
| Education level at baseline |  |  |  | <0.001 |
| Less than high school | 36 (3.63) | 11 (2.31) | 25 (4.90) |  |
| High school degree | 148 (14.90) | 57 (11.95) | 91 (17.84) |  |
| Some college | 312 (31.42) | 141 (29.56) | 170 (33.33) |  |
| College degree | 213 (21.45) | 127 (26.62) | 85 (16.67) |  |
| Post-college degree | 284 (28.60) | 141 (29.56) | 139 (27.25) |  |
| Smoking status |  |  |  | 0.035 |

|  |  |  |  |
| --- | --- | --- | --- |
| Never | 591 (60.12) | 299 (63.62) | 290 (57.20) |
| Former | 333 (33.88) | 151 (32.13) | 179 (35.31) |
| Current | 59 (6.00) | 20 (4.26) | 38 (7.50) |

<sup>a</sup>: Kruskal-Wallis test
<sup>b</sup>: T-test
<sup>c</sup>: Chi-square test
Hypertension: based on definition of SBP ≥ 140, DBP ≥ 90, or current use of anti-hypertensive medication
Missing: hypertension, n = 6; BMI, n = 13; physical activity, n = 51; low-density lipoprotein cholesterol, n = 10; difficulty paying for basics, n = 30; education, n = 6; smoking status, n = 16

More than half (51.5%) of participants were hypertensive, and 4.1% had TRH. Women with HTN were more likely to be Black or Hispanic, to experience economic strain, and have lower levels of education as compared to women without HTN. Women with HTN also had a higher BMI (31.6±7.2 kg/m^2^) as compared to women without HTN (26.8±5.9 kg/m^2^, p<0.001).

The median aldosterone level was 6.79 ng/dL (IQR: 2.74-10.55 ng/dL), but as shown in **Table 2**, women with HTN had significantly higher median aldosterone levels than women without HTN (7.08 [IQR 3.12-10.86] ng/dL versus 6.38 [IQR 2.63-10.13] ng/dL, respectively, p=0.024). Women with TRH had higher median aldosterone levels than women without TRH (8.74 [IQR 5.04-11.67] ng/dL versus 6.75 [IQR 2.66-10.49] ng/dL), but the difference was not statistically significant, potentially due to the small sample of women with TRH (n=41). There were no statistically significant associations between aldosterone and race and ethnicity, economic strain, and education level. However, there was a significant difference in median aldosterone by site (p<0.001). **Figure S1** shows the distribution of serum aldosterone values by race and ethnicity.

**Table 2.** Aldosterone levels (median, IQR) by demographic characteristics.

| Characteristic | Median (IQR) | p-value<sup>a</sup> |
| --- | --- | --- |
| Race/ethnicity |  | 0.097 |
| Black | 6.25 (1.50-10.38) |  |
| White | 6.74 (3.19-11.09) |  |
| Chinese | 6.80 (5.19-10.00) |  |
| Hispanic | 8.08 (5.46-10.41) |  |
| Japanese | 6.73 (1.85-10.07) |  |
| Study site |  | <0.001* |
| Michigan | 3.97 (0.90-9.40) |  |
| Boston | 6.49 (2.61-9.52) |  |
| Chicago | 9.06 (4.95-14.04) |  |
| Davis | 6.95 (4.62-10.49) |  |
| Los Angeles | 6.81 (2.30-10.62) |  |
| New Jersey | 8.08 (5.45-10.69) |  |
| Pittsburgh | 5.86 (2.67-8.63) |  |
| Economic strain |  | 0.069 |
| Very | 3.87 (0.70-8.46) |  |
| Somewhat | 6.88 (2.86-10.47) |  |
| Not at all | 6.87 (3.17-10.58) |  |
| Education level |  | 0.063 |
| Less than high school | 7.21 (5.39-10.13) |  |
| High school degree | 5.81 (1.58-10.01) |  |
| Some college | 6.62 (2.00-10.61) |  |
| College degree | 6.63 (2.84-10.34) |  |
| Post-college degree | 7.18 (4.20-11.68) |  |
| Treatment-resistant hypertension (n=41) |  | 0.131 |
| No | 6.75 (2.66-10.49) |  |
| Yes | 8.74 (5.04-11.67) |  |

<sup>a</sup>: Kruskal-Wallis test
\*: Significant pairwise comparisons based on Dunn's test: Chicago vs. Michigan, Boston, Los Angeles, Pittsburgh; Michigan vs. Davis, Los Angeles, New Jersey; Pittsburgh vs. New Jersey

**Table 3** presents the adjusted associations between aldosterone and HTN and TRH. Higher aldosterone levels were associated with greater odds of HTN; each 1 ng/dL higher aldosterone was associated with 4% greater odds of HTN (95%CI:1.02,1.06; p< 0.001). This finding was consistent in models adjusting for age, race and ethnicity, BMI, smoking status, physical activity, and LDL-c. Models adjust for site instead of race and ethnicity produced similar results. The positive relationship between aldosterone and odds of HTN was also observed in models where aldosterone levels were log-transformed (**Table 3**). Higher aldosterone levels were weakly associated with greater odds of TRH, but this association was not statistically significant (OR=1.04; 95% CI 0.99,1.08; p=0.094).

**Table 3.**
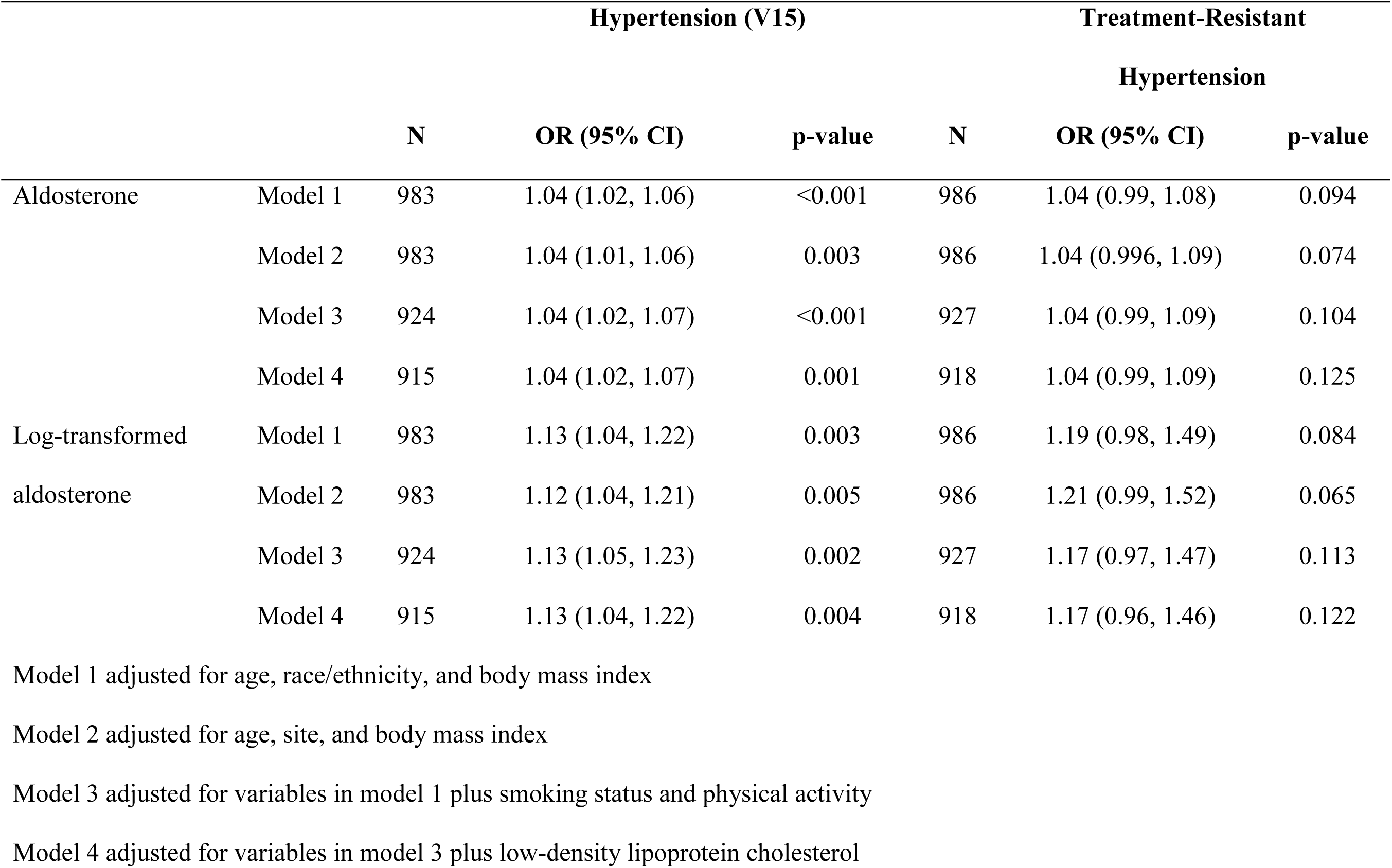
Adjusted logistic regressions of hypertension and treatment-resistant hypertension on aldosterone.

The results demonstrated a statistically-significant interaction between aldosterone and use of anti-hypertensive medication on the relationship between aldolsterone and blood pressure. As shown in **Figure 1**, higher aldosterone was associated with lower systolic blood pressure and pulse pressure among those using anti-hypertensive medications (n=406). The association between each 1 ng/dL higher aldosterone and systolic blood pressure was −0.45 mmHg lower (95% CI −0.74,−0.15; p=0.003) for antihypertensive medication users as compared to non-users (**Table S1**). Specifically, among those using anti-hypertensive medications, women in the 75^th^ percentile of aldosterone levels had a 2.89 mmHg lower systolic blood pressure as compared to women in the 25^th^ percentile of aldosterone (p<0.001), whereas among those not using anti-hypertensive medications, it was 0.62 mmHg higher. Among those not using anti-hypertensive medications, aldosterone was positively associated with systolic and diastolic blood pressure and pulse pressure but the association was not statistically significant.

**Figure 1.**
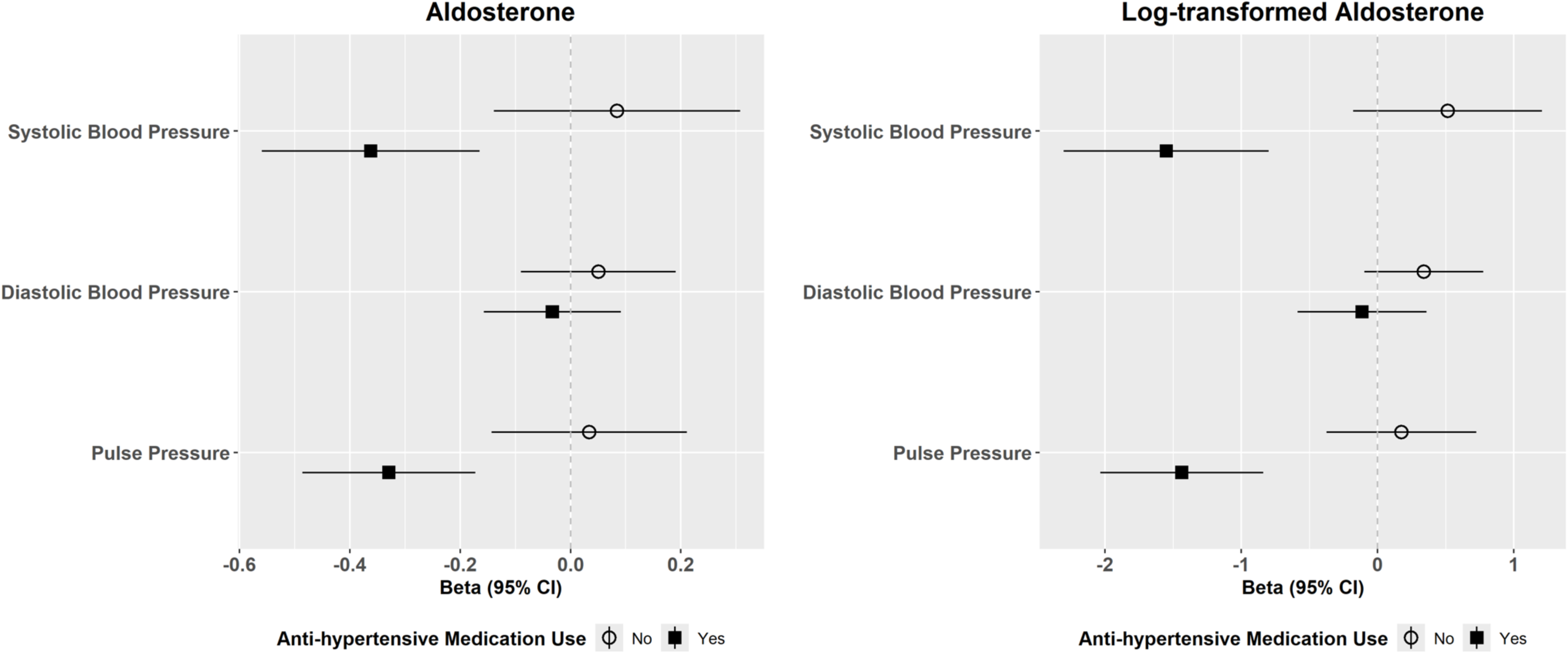
Effect modification by anti-hypertensive medication use of the relationship between aldosterone and systolic blood pressure, diastolic blood pressure, and pulse pressure (N = 871).

## DISCUSSION

The major novel finding of this study is that aldosterone—even within a range commonly considered normal--is associated with the presence of hypertension in post-menopausal women, even after adjusting for important covariates including age, race and ethnicity, body mass index, smoking, physical activity and LDL-c. However, our findings demonstrate important effect modification of the aldosterone and systolic blood pressure relationship by anti-hypertensive medication use. Among those using anti-hypertensive medications, higher aldosterone is associated with lower systolic blood pressure and pulse pressure.

Aldosterone exerts several deleterious effects on cardiac and vascular health, including endothelial dysfunction, cardiac hypertrophy, and inflammation.^13^ Chronic elevation of blood pressure increases cardiac afterload, and aldosterone directly stimulates cardiomyocyte growth and myocardial fibrosis through mineralocorticoid receptor activation. At the vascular level, aldosterone damages the vasculature through multiple pathways.^13^ These molecular derangements impair vasodilation and accelerate vascular remodeling, providing biological mechanisms through which aldosterone elevation—even within the normal range—can drive blood pressure increases in aging women.^14^ In the Framingham Offspring Study, each quartile increase in serum aldosterone among nonhypertensive participants conferred a 16% increased risk of blood pressure elevation and 17% increased risk of incident hypertension over 4 years.^15^ Brown et al. showed that among normotensive Multi-Ethnic Study of Atherosclerosis (MESA) participants with suppressed renin, higher aldosterone levels—even within the normal range (mean 11–12 ng/dL)—predicted incident hypertension (HR 1.18 per 100 pmol/L, 95% CI 1.03–1.36), whereas no such association existed when renin was unsuppressed.^16^ Our findings in postmenopausal women extend these observations, demonstrating that the aldosterone-hypertension relationship persists in an older, exclusively female population.

Among women, the relationship between aldosterone and cardiovascular disease appears particularly significant. In the Framingham cohort, serum aldosterone levels were higher in women and correlated with left ventricular wall thickness in women only—no associations were found in men.^17^ Secondary analysis of the TOPCAT trial revealed that spironolactone was associated with reduced all-cause mortality in women (HR: 0.66, p=0.01) but not in men.^18^ These sex differences potentially could be understood through estrogen receptor-mineralocorticoid receptor crosstalk. Estrogen-activated ERα directly inhibits MR-mediated gene transcription, potentially contributing to premenopausal cardiovascular protection.^19^ Loss of this estrogen-mediated MR inhibition in postmenopause may unmask aldosterone’s deleterious effects, contributing to accelerated cardiovascular risk in aging women. Our finding of an inverse aldosterone-blood pressure relationship among treated women likely reflects treatment dynamics: diuretics and vasodilators – which are first-line treatments for HTN – stimulate compensatory aldosterone production through RAAS activation.^20^ Prior research from SWAN showed increased blood pressure in women across the midlife is driven largely by the timing of menopause,^4^ following which others have shown that women have increased blood pressure lowering during treatment with a mineralocorticoid receptor antagonists (MRA).^21^ In other populations, anti-hypertensive medications, particularly MRAs, a class of diuretics, may have a greater effect on blood pressure in those with primary aldosteronism (PA) versus those without.^22–24^ In a small clinical trial (n=90) of hypertensive patients published by Olivieri and colleagues, a high aldosterone-to-renin ratio did not significantly predict response to an MRA. Notably, most women in the Olivieri study were postmenopausal, and compared to men, showed nearly 2-fold greater SBP reduction at 2 months (16.4 vs. 8.2 mmHg)—suggesting that postmenopausal women may be particularly responsive to MRA therapy. This enhanced treatment response in women with aldosterone-mediated hypertension provides a plausible explanation for our observation that higher aldosterone levels associate with lower blood pressure among treated women: those with the most aldosterone-driven hypertension may achieve the greatest blood pressure reductions when adequately treated with medications that block or stimulate compensatory aldosterone pathways.

Despite the strengths in our study including a large, well-characterized cohort, availability of information about important confounding measures including age, race/ethnicity, body mass index, smoking, and physical activity, and information about anti-hypertension medication use, there are some important limitations to acknowledge. Clinical definitions of PA require information of both aldosterone and renin levels, and we do not have plasma renin values available in our cohort. Further, while we postulate that our findings of effect modification by antihypertensive medication use may be explained by increased activity of the RAAS system, we could not restrict this analysis to the small number of treatment naïve women with HTN. Data from SWAN suggests that, among women taking an anti-hypertensive medication, more than half received a diuretic medication, which is consistent with our interpretation of the data.

Our findings demonstrate that subclinical aldosterone elevation associates with hypertension in postmenopausal women, challenging the notion that “normal” aldosterone levels are necessarily healthy. These results suggest that sex- and age-specific aldosterone ranges may be needed to guide clinical decision-making and that targeted mineralocorticoid receptor antagonism or dietary sodium modification warrants investigation for mitigating the blood pressure rise characteristic of the menopausal transition.

## Data Availability

The data that support the findings of this study are available from the senior author upon reasonable request. Data will be made available through a data use agreement process with the Study of Women’s Health Across the Nation.

## Acknowledgements

The Study of Women’s Health Across the Nation (SWAN) has grant support from the National Institutes of Health (NIH), DHHS, through the National Institute on Aging (NIA), the National Institute of Nursing Research (NINR) and the NIH Office of Research on Women’s Health (ORWH) (Grants U01NR004061; U01AG012505, U01AG012535, U01AG012531, U01AG012539, U01AG012546, U01AG012553, U01AG012554, U01AG012495, and U19AG063720). The content of this manuscript is solely the responsibility of the authors and does not necessarily represent the official views of the NIA, NINR, ORWH or the NIH.

## Clinical Centers

*University of Michigan, Ann Arbor – Carrie Karvonen-Gutierrez, PI 2021 –present, Siobán Harlow, PI 2011 – 2021, MaryFran Sowers, PI 1994-2011*; *Massachusetts General Hospital, Boston, MA – Sherri-Ann Burnett-Bowie, PI 2020 – Present; Joel Finkelstein, PI 1999 – 2020*; *Robert Neer, PI 1994 – 1999; Rush University, Rush University Medical Center, Chicago, IL – Imke Janssen, PI 2020 – Present; Howard Kravitz, PI 2009 – 2020*; *Lynda Powell, PI 1994 – 2009; University of California, Davis/Kaiser – Elaine Waetjen and Monique Hedderson, PIs 2020 – Present; Ellen Gold, PI 1994 - 2020*; *University of California, Los Angeles – Arun Karlamangla, PI 2020 – Present; Gail Greendale, PI 1994 - 2020*; *Albert Einstein College of Medicine, Bronx, NY – Carol Derby, PI 2011 – present, Rachel Wildman, PI 2010 – 2011; Nanette Santoro, PI 2004 – 2010; University of Medicine and Dentistry – New Jersey Medical School, Newark – Gerson Weiss, PI 1994 – 2004;* and the *University of Pittsburgh, Pittsburgh, PA – Rebecca Thurston, PI 2020 – Present; Karen Matthews, PI 1994 - 2020*.

## NIH Program Office

*National Institute on Aging, Bethesda, MD – Rosaly Correa-de-Araujo 2020 - present; Chhanda Dutta 2016 - present; Winifred Rossi 2012–2016; Sherry Sherman 1994 – 2012; Marcia Ory 1994 – 2001; National Institute of Nursing Research, Bethesda, MD – Program Officers*.

## Central Laboratory

*University of Michigan, Ann Arbor – Daniel McConnell* (Central Ligand Assay Satellite Services).

## NIA Biorepository

*Rosaly Correa-de-Araujo 2019 - Present*; SWAN Repository: *University of Michigan, Ann Arbor – Siobán Harlow 2013 - 2018; Dan McConnell 2011 - 2013; MaryFran Sowers 2000 – 2011*.

## Coordinating Center

*University of Pittsburgh, Pittsburgh, PA – Maria Mori Brooks, PI 2012 -present; Kim Sutton-Tyrrell, PI 2001 – 2012; New England Research Institutes, Watertown, MA - Sonja McKinlay, PI 1995 – 2001*.

Steering Committee: Susan Johnson, Current Chair

Chris Gallagher, Former Chair

## We thank the study staff at each site and all the women who participated in SWAN. Sources of Funding

This work was supported by the National Institutes of Health (NHLBI) R01 award R01HL164678.

## SUPPLEMENTAL MATERIAL

- Figure S1
- Supplemental Methods

## Nonstandard Abbreviations and Acronyms

SWAN: Study of Women’s Health Across the Nation
NHANES: National Health and Nutrition Examination Survey
RAAS: renin-angiotensin-aldosterone system
ALDO: aldosterone
LLOQ: Lower limit of quantification
MT: Menopausal transition
TRH: Treatment-resistant hypertension
TOPCAT: Treatment of Preserved Cardiac Function Heart Failure with an Aldosterone Antagonist

## Novelty and Relevance

### What Is New?

- In a large, well-characterized longitudinal cohort of postmenopausal women, serum aldosterone—measured by LC-MS/MS across a range that includes levels conventionally regarded as normal—is independently associated with prevalent hypertension after adjustment for age, race/ethnicity, body mass index, smoking, physical activity, and LDL cholesterol.
- · Among women using antihypertensive medications, higher aldosterone is associated with lower systolic blood pressure and pulse pressure, demonstrating significant effect modification by antihypertensive medication use on the aldosterone–blood pressure relationship.
- · Treatment-resistant hypertension was associated with higher aldosterone levels, though this association did not reach statistical significance, likely reflecting limited statistical power given the small number of affected women.

### What Is Relevant?

- These findings extend evidence linking subclinical aldosterone excess to hypertension risk into an older, exclusively female population, a group underrepresented in prior aldosterone research.
- The inverse association between aldosterone and blood pressure among treated women is consistent with compensatory RAAS activation by diuretics and vasodilators, and raises the possibility that women with the highest aldosterone burden achieve the greatest blood pressure reduction from treatments that block mineralocorticoid pathways.
- The results support investigation of sex- and age-specific aldosterone reference ranges and targeted mineralocorticoid receptor antagonism as a strategy to address the blood pressure rise characteristic of the postmenopausal period.

### Clinical/Pathophysiological Implications

- Aldosterone levels within the conventionally accepted normal range are associated with hypertension in postmenopausal women, suggesting that current reference intervals may not adequately capture clinically relevant aldosterone excess in this population and that sex- and age-specific thresholds warrant formal evaluation.
- The significant interaction between aldosterone and antihypertensive medication use on systolic blood pressure is consistent with a pathophysiological model in which first-line antihypertensive agents—particularly diuretics—drive compensatory RAAS activation, elevating aldosterone in treated women and potentially sustaining or worsening aldosterone-mediated end-organ effects despite apparent blood pressure control.
- Loss of estrogen-mediated mineralocorticoid receptor inhibition after menopause may unmask aldosterone’s pressor and fibrotic effects, providing a mechanistic rationale for the excess cardiovascular risk that accumulates in postmenopausal women and for the observed sex differences in response to mineralocorticoid receptor antagonism.
- The association between higher aldosterone and greater odds of hypertension—combined with prior evidence that postmenopausal women exhibit an enhanced blood pressure response to mineralocorticoid receptor antagonists—supports prospective evaluation of targeted mineralocorticoid receptor antagonism as a blood pressure-lowering strategy specifically in postmenopausal women with elevated aldosterone.
- The subclinical aldosterone excess identified here, in the absence of renin suppression data sufficient to diagnose primary aldosteronism, highlights the clinical relevance of the broader spectrum of autonomous aldosterone production and argues for systematic aldosterone assessment in postmenopausal women with incident or poorly controlled hypertension.

